# Epigenetic and Structural Brain Aging and their Associations with Major Depressive Disorder and Inflammatory Markers

**DOI:** 10.1101/2024.11.21.24317719

**Authors:** Eileen Y. Xu, Claire Green, Daniel L. McCartney, Laura K.M. Han, Kathryn L. Evans, Rosie M. Walker, Danni A. Gadd, Douglas Steele, Gordon Waiter, Archie Campbell, Stephen M. Lawrie, James H. Cole, Andrew M. McIntosh, Xueyi Shen, Heather C. Whalley

## Abstract

**Background:** A growing body of evidence suggests that Major Depressive Disorder (MDD) may be associated with premature biological aging. However, most studies have examined brain-based and DNAm-based measures of biological age (BioAge) in isolation. Previous studies also suggest the relevance of inflammation, yet the relationship between MDD, BioAge and inflammation remains unclear.

**Method:** We investigated two well-studied BioAge measures: BrainAge and DNA methylation age (DNAmAge) in Generation Scotland (GS:STRADL; BrainAge N=1,067; DNAmAge N=684; 26-76 years) and UK Biobank (UKB, BrainAge N=12,018, 45-80 years). Premature brain and DNAm aging was operationalised as ‘Predicted Age Difference’ (Brain-PAD and DNAm-PAD, respectively). We tested individual and additive contributions of Brain-PAD and DNAm-PAD to lifetime/current MDD using logistic regression, followed by exploratory analyses of acute inflammatory biomarkers as mediators of this relationship.

**Results:** Lifetime MDD cases showed significantly higher BrainAge and DNAmAge, ranging from 1.60-2.45 years increase compared to controls; no differences were found for DNAmAge_Horvath_ or for BrainAge in UKB. Lifetime MDD associated with DNAm-PAD_GrimAge_, DNAm-PAD_PhenoAge_ and Brain-PAD, ranging from β = .22 - .27 (UKB Brain-PAD β = .05). DNAm-PAD and Brain-PAD demonstrated shared and distinctive contributions to lifetime MDD (DNAm-PAD_PhenoAge_ plus Brain-PAD explained maximum variance, AUC=0.69, R^2^=9%). Six inflammation biomarkers associated with current, but not lifetime MDD; no significant mediation effects were found.

**Conclusions:** Our findings highlight shared and distinct contributions of premature brain and DNAm aging in lifetime MDD. We found no evidence for a mediating role of inflammation, however future work utilizing more stable biomarkers may elucidate potential biological mechanisms.

## Introduction

Major Depressive Disorder (MDD) is the leading cause of disability worldwide(1, 2). MDD is comorbid with a number of age-related diseases and phenotypes such as type 2 diabetes, cognitive decline, dementia, cardiovascular diseases and stroke(3). In addition, MDD is associated with an increased risk of mortality, which persists up to two decades after a depressive episode(4). This has informed the theory that MDD may be a state of premature biological aging, where physical and psychological stressors trigger biochemical alterations which then lead to age-related changes at the molecular level(5).

Two popular estimators of biological age (BioAge) include those based on DNA methylation (DNAmAge), and on structural neuroimaging data (BrainAge)(6–11). DNAm at certain CpG sites has been associated with chronological age(12, 13), resulting in the development of several DNAmAge estimators (“DNAm clocks” or “epigenetic clocks”) that can predict mortality and diseases(6, 8, 9, 13–17). Similarly, brain structures are well-known to change over lifecourse, and estimations of BrainAge are a promising tool that can be used to quantify compromised brain health in the context of neurobiological disorders(11). Estimates of BioAge (from DNAm and brain imaging based) are then either subtracted from, or regressed on, chronological age to derive a biologically informative summary score (the ‘Predicted Age Difference’, or PAD) which reflects deviation from normal aging trajectories, where positive PAD indicates premature biological aging.

Premature biological aging in MDD has been indicated previously through separate investigations of both DNAmAge and BrainAge measures. For example, an increase in DNAmAge compared to chronological age (positive DNAm-PAD) has been shown to associate with a range of diseases comorbid with MDD, such as cardiovascular diseases, and in smaller studies of MDD itself(7, 15–19). However, it is less clear whether DNAm-PAD is directly involved in the etiology of MDD. With regard to brain aging, studies on the association between Brain-PAD and MDD have also indicated premature brain aging in a wide range of age groups, including older and midlife participants to young adults and adolescents(20–22). MDD is also phenotypically and genetically correlated with disorders that have established associations with premature brain aging, such as Alzheimer’s and cardiovascular diseases(7, 15–18). Thus, while DNAm-PAD and Brain-PAD have previously been independently associated with MDD, combining the two types of aging measures could potentially boost predictive power and aid understanding of risk factors(*23, 24*). Concurrent investigation of peripheral and brain-specific PAD measures may also facilitate understanding of shared and distinct mechanisms of premature aging in MDD. In terms of underlying biological mechanisms, premature aging has consistently been associated with inflammation(*25*). While numerous studies have demonstrated that the pathogenesis of MDD also involves inflammation(*26*),(*27*), the extent to which acute and chronic inflammation contributes to the relationship between MDD and premature biological aging remains uncertain.

The lack of an integrated investigation into the role of different biological aging markers in MDD, and the relationship between inflammation, biological aging and MDD, is likely due to the lack of available multi-modal biological samples collected in the same cohort at the same timepoint. In the present study, we therefore used a population-based cohort of N=658 unrelated individuals with DNAm, structural MRI and inflammatory markers all available from the same data collection point, with a second brain imaging replication cohort of N=∼20,000 participants from UK Biobank. We first tested whether DNAm-PAD and Brain-PAD were higher in MDD cases versus controls, before conducting replication analysis of Brain-PAD in the independent UKB replication sample. We then tested the unique contribution of each of these biomarkers of aging to MDD. Finally, we used structural equational modelling to explore the role of inflammatory markers in mediating the association between BioAge-PAD and MDD.

## Methods and Materials

### Generation Scotland

Generation Scotland is a family- and community-based population cohort. A total of ∼20,000 participants were recruited from across Scotland(28). A subset of participants attended brain MRI scanning in the Generation Scotland: Stratifying Resilience and Depression Longitudinally follow-up study (GS:STRADL)(29, 30); N = 1,067 individuals (60 % Female; age M (SD) = 59.8 (10.1) years) with brain MRI data were included in the current study. DNAm data was collected concurrently with imaging clinic visits and DNAmAge analyses included N = 684 individuals (56.3% Female; age M (SD) = 60.5 (9.2) years).

Overlap between these two samples included N = 628 unrelated participants that both neuroimaging and DNAm data collected (57.2% Female; age M (SD) = 60.5 (9.3) years). Details of recruitment and cohort profiling can be found elsewhere(28). Written consent was obtained from all participants. The study was approved by the NHS Tayside Research Ethics committee (05/s1401/89).

### UK Biobank

UK Biobank is a population cohort study of half a million mid- to late-life participants, recruited from across the United Kingdom. A subset of UK Biobank neuroimaging data (released in 2019)(31), was used in the present study as a replication dataset for Brain Age analysis. Written consent was obtained for all participants. Withdrawn participants up until completion of the study were not included in the analysis. Ethical permission was obtained through the National Health Service (NHS) Research Ethics Service (11/NW/0382, UK Biobank project #4844).

The neuroimaging sample from UK Biobank used for replication analysis in the current study comprised N = 12,018 unrelated participants with complete lifetime MDD data (53.3% Female; age M (SD) = 63.2 (7.4) years).

Descriptive statistics for each of these samples are included in Table 1, Supplementary Table S1 and Figures S2-S3.

**Table 1:**
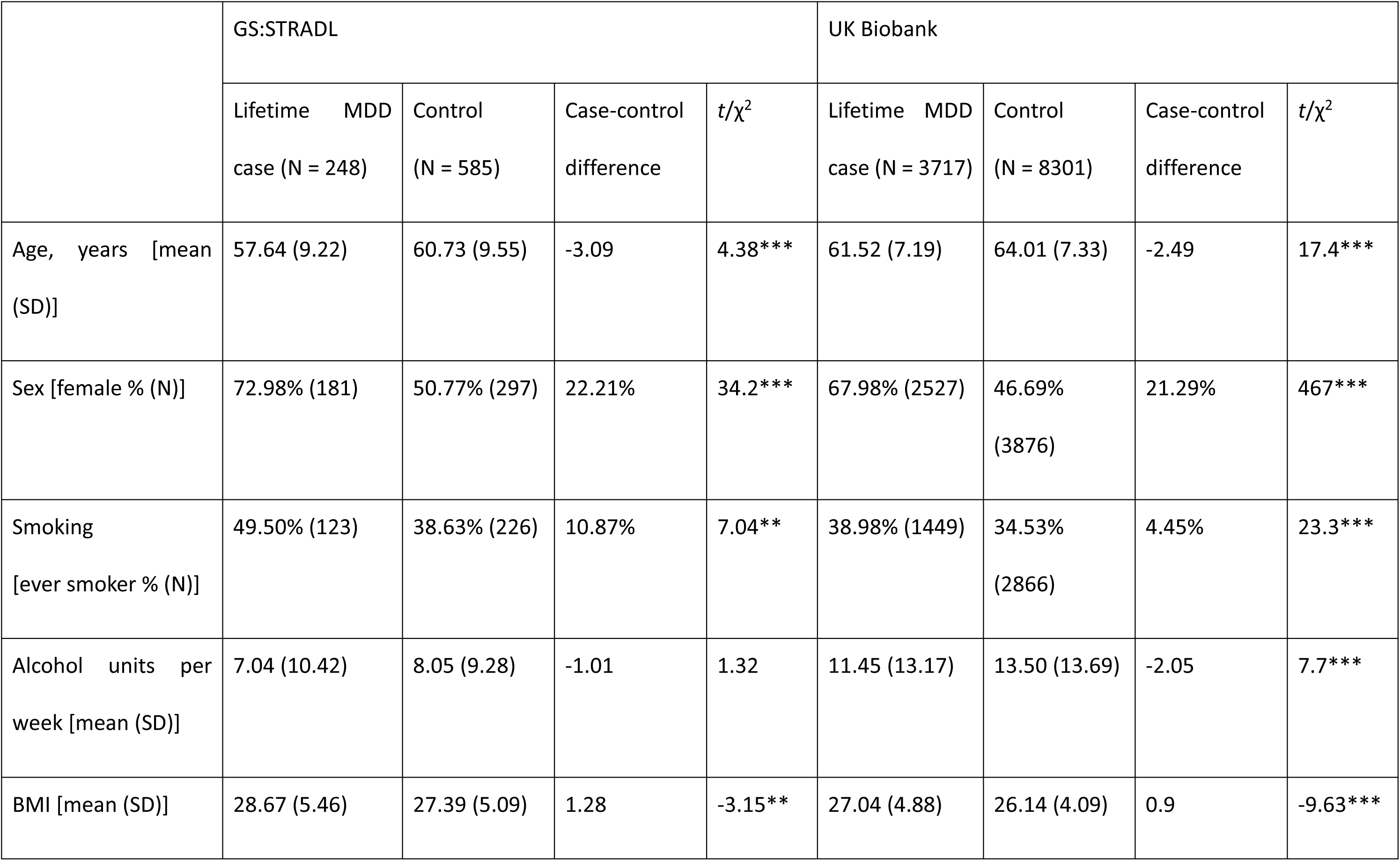
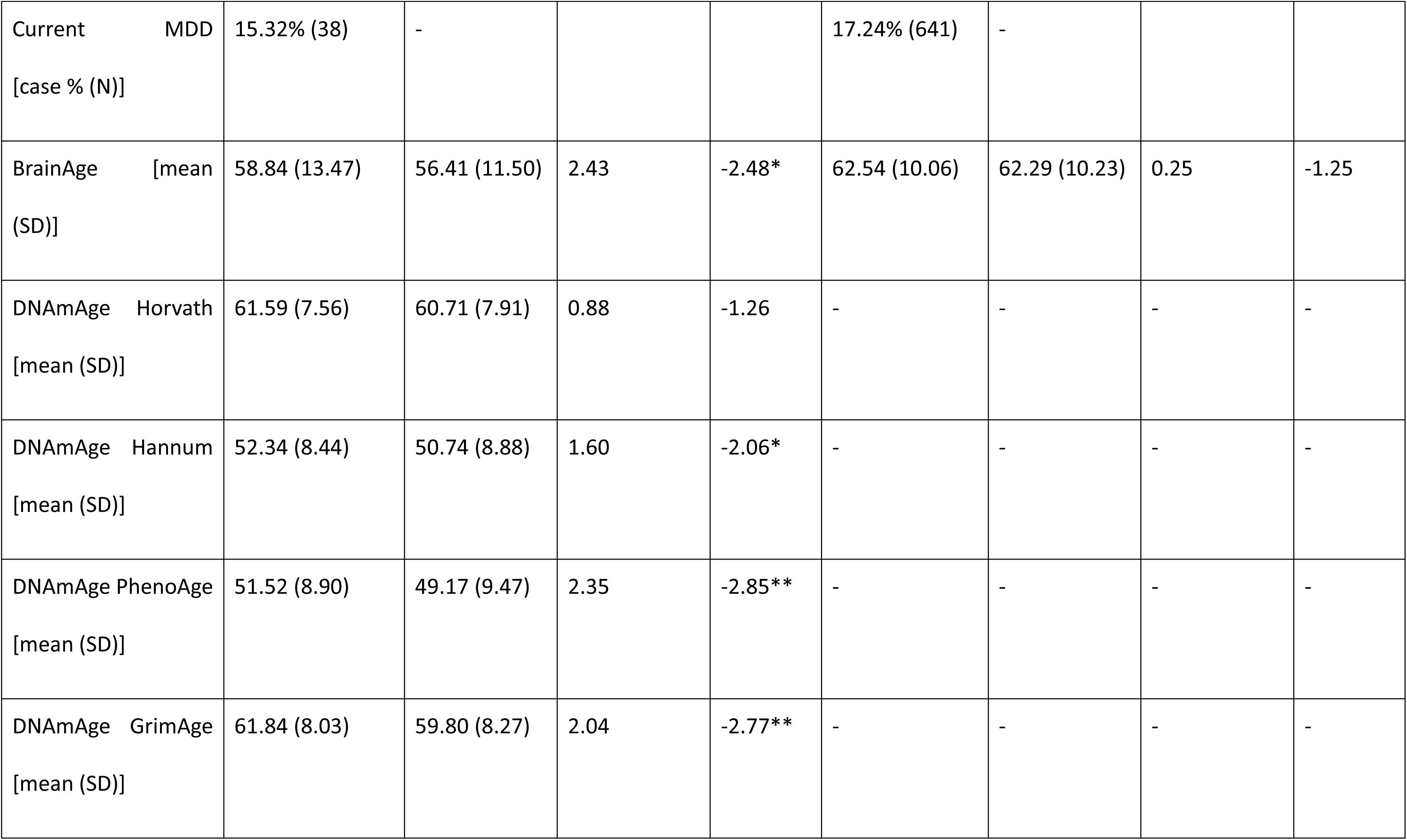
Demographic information for lifetime MDD cases and controls in GS:STRADL and UK Biobank. BMI = body mass index. GS:STRADL = Generation Scotland: Stratifying Resilience and Depression Longitudinally. *p<0.05; **p<0.01; ***p<0.001.

### Definition of MDD

Participants in GS:STRADL were assessed for MDD history using the Structured Clinical Interview for DSM-IV disorders. Diagnostic criteria in this assessment are consistent with symptom criteria within the Diagnostic and Statistical Manual of Mental Disorders 4^th^ edition (DSM-IV)(30). Participants were classed as lifetime MDD cases or controls based on whether they had experienced at least one major depressive episode by the time of assessment. Current MDD and severe MDD cases and controls were also separated from lifetime MDD cases using the SCID. Finally, an antidepressant use phenotype (antidepressant MDD) was determined using participant self-reported medication usage at the time of interview and supported by data from linked health records where available.

In UK Biobank, lifetime MDD cases and controls were identified based on the self-report Composite International Diagnostic Interview – Short Form (CIDI-SF), introduced as an online questionnaire after the baseline assessment(32). Current MDD cases were classified from lifetime MDD cases based on responses to items from the 9-item Patient Health Questionnaire (PHQ-9) which assessed MDD symptoms in the preceding 2 weeks(33, 34). Severe MDD cases were identified based on the ‘probable’ MDD definition, which considers self-reported history of hospital admission and self-reported depressive symptoms(34, 35). Finally, antidepressant MDD cases were identified based on self-reported prescribed medications being taken at the time.

Results for lifetime MDD are reported as the main findings in this manuscript. Additional findings for current, severe MDD and antidepressant use phenotypes are reported in supplementary materials (Figures S2-S5 and Tables S2-S4).

### DNAm-PAD

DNAm data from GS:STRADL were profiled from whole blood samples using the Illumina Infinium MethylationEPIC BeadChip (Illumina Inc., San Diego, California) according to the manufacturer’s protocol. Details for data preprocessing have been published elsewhere(36). DNA methylation was measured in the GS:STRADL samples in two sets. Quality control and pre-processing were performed for each set of samples separately in R 3.6.1(37) using minfi(38) and wateRmelon(39), removing samples where >0.5% of CpGs had a detection p-value >0.01, probes where >1% of samples had a detection p-value >0.01 and probes with a beadcount of <3 in >5% of samples. Data were background-corrected using normal-exponential out-of-band (“noob”) pre-processing(40). Finally, reported biological sex assigned at birth was confirmed by DNAm data and no further exclusions were made, leaving a total N=685 GS:STRADL samples for analysis.

DNAmAge was calculated from GS:STRADL DNAm data using the DNA Methylation Age Calculator online resource (http://dnamage.genetics.ucla.edu/)(9, 41). A total of four DNAmAge measures were generated: Horvath age, Hannum age, PhenoAge and GrimAge(6, 8, 9, 14). More details on each DNAmAge measure are presented in the Supplementary Materials.

DNAm-PAD was defined as the residual term from regressing each DNAmAge estimate against chronological age, including sex, methylation processing set, and DNAm-estimated cell proportions taken at blood draw as covariates in the same model. Following this, we rescaled DNAm-PAD residuals to the original DNAmAge mean and standard deviation for each DNAmAge measure to generate an estimate of DNAmAge with the effects of chronological age and covariates removed, in units of years. Throughout the manuscript we refer to estimate of DNAmAge for each epigenetic clock as DNAmAge_clock_, with units of years; DNAm-PAD_clock_ refers to the residual term itself whereby a positive DNAm-PAD indicates accelerated, or premature aging and a negative DNAm-PAD indicates decelerated, or delayed aging.

### Brain-PAD

GS:STRADL MRI data was acquired and pre-processed using a unified protocol(30). In brief, data was acquired at two sites using a 3T Philips (Amsterdam, The Netherlands) Achieva TX scanner and 32-channel head coil at one and a 3T Siemens (Munich, Germany) Prisma-FIT scanner and 20-channel head coil at the other. T1-weighted images from UK Biobank (released in 2019) was acquired at two sites, each using a 3T Siemens Skyra scanner and 32-channel head coil. Recruitment and raw T1-weighted image acquisition were completed by the UK Biobank imaging team(31). Further details of acquisition protocols have been published previously for both GS:STRADL(30) and UK Biobank(31).

BrainAge was predicted using the software package ‘brainageR’, version 2.1 (https://github.com/james-cole/brainageR)(10). Preprocessing steps in the brainageR packages included SPM12 (https://www.fil.ion.ucl.ac.uk/spm/doc/) and FSL. BrainageR uses principal components derived from voxel-wise volumetric data in normalised grey matter, white matter and CSF segmentations to predict age. The brainageR model was trained on 3,377 healthy participants from 7 sites, aged 18-92 years(10). Similar to DNAm-PAD, we defined Brain-PAD as the residual term from regressing BrainAge against chronological age with sex and scan site covariates. In UK Biobank replication analyses, scanner head position coordinates included in this model to adjust for potential bias caused by uneven static MRI field. Following this, Brain-PAD residuals were rescaled to the original BrainAge mean and standard deviation to generate a new estimate of BrainAge in years, minus the effects of chronological age. Throughout the manuscript, BrainAge refers to the above estimated age in years; Brain-PAD refers to the age difference relative to chronological age – positive Brain-PAD indicates accelerated or premature aging, negative Brain-PAD indicates delayed aging.

### Inflammatory markers

Inflammatory markers in GS:STRADL were profiled from whole blood samples collected along with the MRI assessment. Blood samples were processed at National Health Service laboratories in Ninewells Hospital and Aberdeen Royal Infirmary. Details of blood sample processing and profiling were stated in a previous study(30).

For brevity, only blood markers significantly associated with MDD are reported in the main text; the full list of the 19 blood markers tested in GS:STRADL can be found in the supplementary materials (Table S2).

### Statistical analysis

Statistical analyses were performed in R version 4.1.0 and 4.1.3(37). Model performance and comparison metrics were generated using the R package “performance”, version 0.10.4(42). In GS:STRADL analyses, p-values were FDR-adjusted across the five BioAge measures. P-values were not adjusted for multiple comparison in UKB due to there being only one BioAge measure (i.e., BrainAge). In all analyses, statistical significance was determined as FDR-adjusted p_FDR_ < 0.05, or α < 0.05 for unadjusted analyses.

### BioAge measures

All five BioAge estimations (four DNAmAge estimates and BrainAge) were validated by comparing against chronological age. Pearson’s correlation was calculated between BioAge measures and chronological age. Case-control differences in mean BioAge were then compared using t-tests.

### MDD case-control differences in BioAge-PAD

Case-control differences in BioAge-PAD were compared using logistic regression, with MDD status as the binary outcome (the ‘glm’ function in R). Associations between MDD status and BioAge-PAD were tested with age and sex covariates included. In Brain-PAD models, assessment centre and scanner head position coordinates (only available in UK Biobank) were also included as covariates. Where there was a significant interaction between BrainAge and assessment centre, this interaction term was added as an additional covariate.

Standardised log-transformed odds ratios are reported as effect sizes. A positive effect size indicates increased aging in cases compared to controls.

### Additive contributions of DNAm-PAD and Brain-PAD

Additive contributions of DNAm-PAD and Brain-PAD were assessed by stepwise comparison of minimally-adjusted models. One null model and two sets of testing models were created to compare the added contribution of Brain-PAD to DNAm-PAD-only models for each DNAm-PAD measure (i.e. tested individually), and vice versa:

1. H1 model: Lifetime MDD ∼ age + sex
2. H2 models: Lifetime MDD ∼ age + sex + DNAm-PAD *<u>OR</u>* Brain-PAD
3. H3 model: Lifetime MDD ∼ age + sex + DNAm-PAD *<u>AND</u>* Brain-PAD

H2 models were compared against the H1 model to evaluate variance explained of MDD by each individual DNAm-PAD or Brain-PAD. The H3 model was then compared against each of the H2 models to obtain the increased variance explained by including both DNAm-PAD and Brain-PAD together.

Area Under the Curve (AUC) and Tjur’s R^2^ were compared for each set of models to quantify improvements in model fit. Tjur’s R^2^ was selected over other pseudo-R^2^s due to its similarity with R^2^ for linear models(43); Tjur’s R^2^ values closer to 1 indicate greater separation between predicted probabilities for cases and controls. Model comparisons were made using Chi-squared tests using the R functions ‘test_performance’ and ‘compare_performance’ from the ‘performance’ package. For completeness, a heatmap of correlations between the BioAge-PAD measures themselves is presented in Supplementary Figure S6.

### Mediation effect of inflammatory markers

To investigate the relationship between MDD, biological aging and peripheral inflammation, we conducted an exploratory mediation analysis using inflammatory markers as mediation variables, DNAm-PAD or Brain-PAD as the predictor variables and MDD as the outcome (Figure 3). The aim was to test if inflammation could explain part of the relationship between biological aging and MDD. We first tested associations between individual inflammatory markers and MDD phenotypes; inflammatory markers associated with either lifetime MDD or current MDD were subsequently tested as potential mediators.

Mediation analysis was performed using ‘lavaan’ R package(44). Significant mediation was determined as both a significant indirect effect and total effect (p<0.05).

## Results

Descriptions of GS:STRADL and UK Biobank samples are reported in Table 1. Participant ages were similar across the two samples. Notably the number of cases with current MDD was relatively small, with N=38 cases in GS: STRADL (15% of MDD cases) and N=641 in UK Biobank (17% of MDD cases).

### BioAge measures

BioAge was highly correlated with chronological age across all DNAmAge estimates, see supplementary Figure S1. DNAmAge_Hannum_ showed the strongest correlation (Pearson’s r = .90; p < .001), followed by DNAmAge_Horvath_ (r = .88; p < .001), DNAmAge_GrimAge_ (r = .86; p < .001), and DNAmAge_PhenoAge_ (r = .83; p < .01). Accuracy of BrainAge predictions in terms of correlation with chronological age, were r = .82 (p < .001) for GS:STRADL and r = .79 (p < .001) for UK Biobank.

Lifetime and current MDD cases had higher mean BioAge compared to controls (Table 1). For lifetime MDD the greatest mean difference was for BrainAge (2.43 years), followed by DNAmAge_PhenoAge_ (2.35 years), DNAmAge_GrimAge_ (2.04 years) and DNAmAge_Hannum_ (1.60 years); the smallest difference was for DNAmAge_Horvath_ (0.88 years). Case-control differences were statistically significant for all except DNAmAge_Horvath_

For current MDD, DNAmAge_GrimAge_ showed the greatest difference between cases and controls (4.69 years), followed by DNAmAge_PhenoAge_ (4.27 years), DNAmAge_Hannum_ (3.96 years), BrainAge (2.79 years) and DNAmAge_Horvath_ (2.20 years). Case-control differences were statistically significant for DNAmAge_GrimAge_ and DNAmAge_PhenoAge_ only.

Smaller case-control differences were found for BrainAge in UKB for lifetime MDD (0.25 years) and current MDD (0.61 years); these were not statistically significant.

### MDD case-control differences in BioAge-PAD

#### Lifetime MDD

Prior to FDR correction, lifetime MDD was positively associated with DNAm-PAD_Hannum_ (β = .20; 95% CI: .02 - .38; p = .03), DNAm-PAD_PhenoAge_ (β = .27; 95% CI: .09 - .46; p < .01) and DNAm-PAD_GrimAge_ (β = .26; 95% CI: .08 - .45; p < .01). No significant associations were found for DNAm-PAD_Horvath_. Brain-PAD was also positively associated with lifetime MDD (β = .22; 95% CI: .06 - .38; p < .01). Following FDR correction for multiple testing, significant associations with MDD persisted for Brain-PAD, DNAm-PAD_PhenoAge_ and DNAm-PAD_GrimAge_. Consistent with the results from GS:STRADL, Brain-PAD in UK Biobank was also positively associated with lifetime MDD in this model (β = .05; 95% CI: .01 - .09; p = .02) (Figure 1).

**Figure 1:**
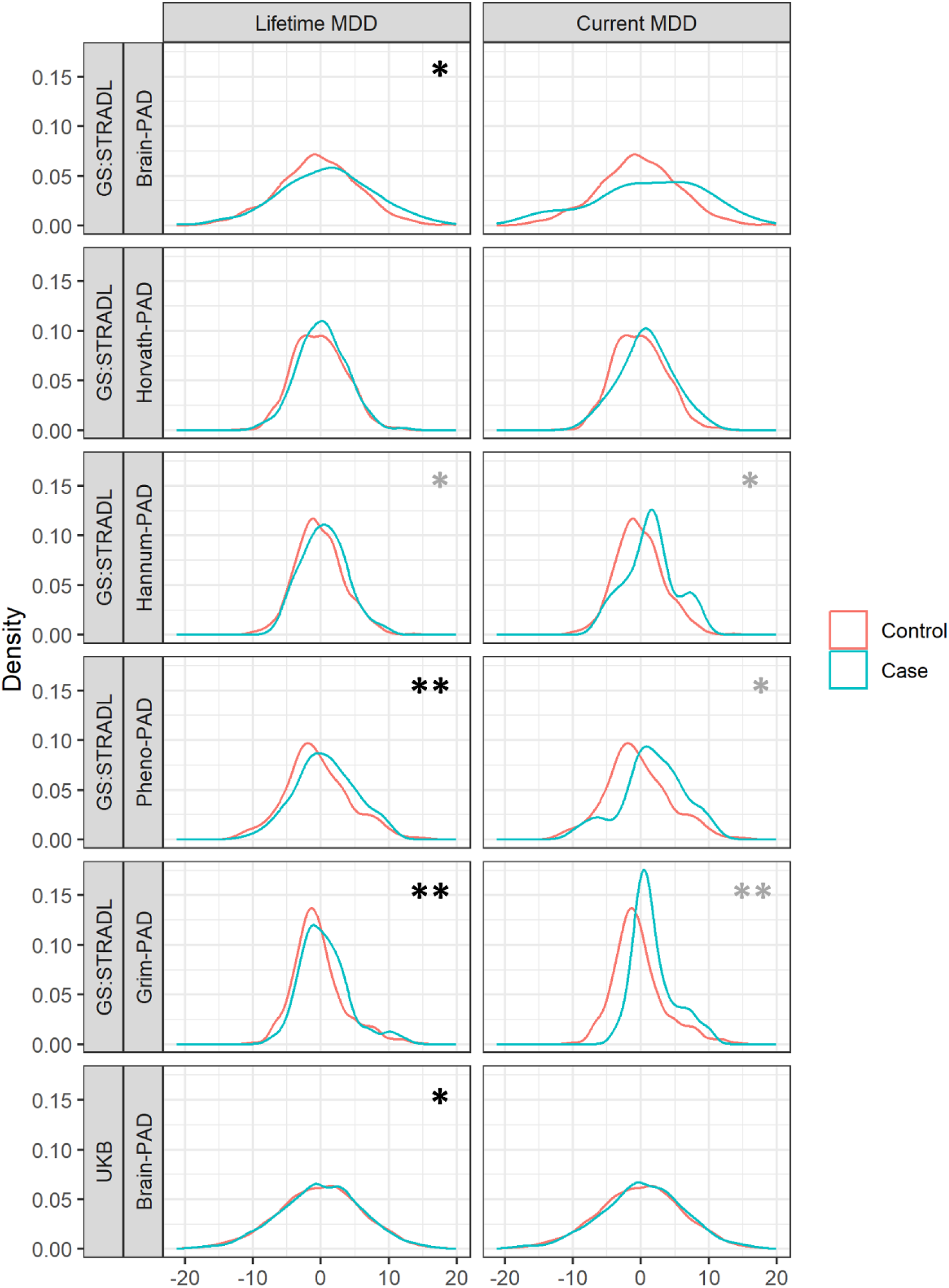
Distribution of BioAge-PAD measures across cases and controls for lifetime and current MDD phenotypes. Grey asterisks represent statistical significance prior to FDR correction, black asterisks represent statistical significance after FDR correction. PAD = Predicted age difference. GS:STRADL = Generation Scotland: Stratifying Resilience and Depression Longitudinally. UKB = UK Biobank. * = p<.05, ** = p<.01, *** = p<.001.

#### Current MDD

Prior to FDR correction, current MDD was associated with DNAm-PAD_Hannum_ (β = .43; 95% CI: .04 - .82; p = .03), DNAm-PAD_PhenoAge_ (β = .42; 95% CI: .02 - .82; p = .04) and DNAm-PAD_GrimAge_ (β = .53; 95% CI: .14 - .90; p < .01). However, none of these associations survived FDR correction. No significant associations were found for DNAm-PAD_Horvath_ or Brain-PAD with current MDD in either GS:STRADL and UK Biobank (Figure 1, right).

### Additive contributions of DNAm-PAD and Brain-PAD

Comparisons were performed to test the combined contributions of DNAm-PAD and Brain-PAD over and above a single measure only (DNAm-PAD or Brain-PAD alone), when predicting MDD status.

For lifetime MDD, the model with greatest R^2^ was for DNAm-PAD_PhenoAge_ and Brain-PAD combined (AUC=0.69, R^2^=9%) (Figure 2B and 2C). The addition of Brain-PAD to DNAm-PAD models significantly improved fit for DNAm-PAD_Horvath_ (χ^2^ = 5.77; p = .02; ΔAUC = .01; ΔR^2^ = .01), DNAm-PAD_Hannum_ (χ^2^ = 5.13; p = .02; ΔAUC = .01; ΔR^2^ = .01), DNAm-PAD_PhenoAge_ (χ^2^ = 4.43; p = .04; ΔAUC = .01; ΔR^2^ = .01) and DNAm-PAD_GrimAge_ (χ^2^ = 4.07; p = .04; ΔAUC = .001; ΔR^2^ = .01) (Figure 2C). The reverse analyses of adding DNAm-PAD to Brain-PAD models also resulted in significant improvement in model fit, with the exception of DNAm-PAD_Horvath_ (Hannum: χ^2^ = 3.99; p = .05, ΔAUC = .007; ΔR^2^ = .012; PhenoAge: χ^2^ = 7.50; p = .006, ΔAUC = .014; ΔR^2^ = .01; GrimAge: χ^2^ = 5.63; p =, ΔAUC = .018; ΔR^2^ = .008) (Figure 2D). When all DNAm-PAD measures were combined, the addition of Brain-PAD slightly improved model fit (χ^2^ = 4.00; p = .045; ΔAUC = .008; ΔR^2^ = .009) (Figure 2A). Similarly, adding all DNAm-PAD measures simultaneously to Brain-PAD also resulted in improved fit (χ^2^ = 10.93; p = .027; ΔAUC = .02; ΔR^2^ = .015) (Figure 2B).

**Figure 2:**
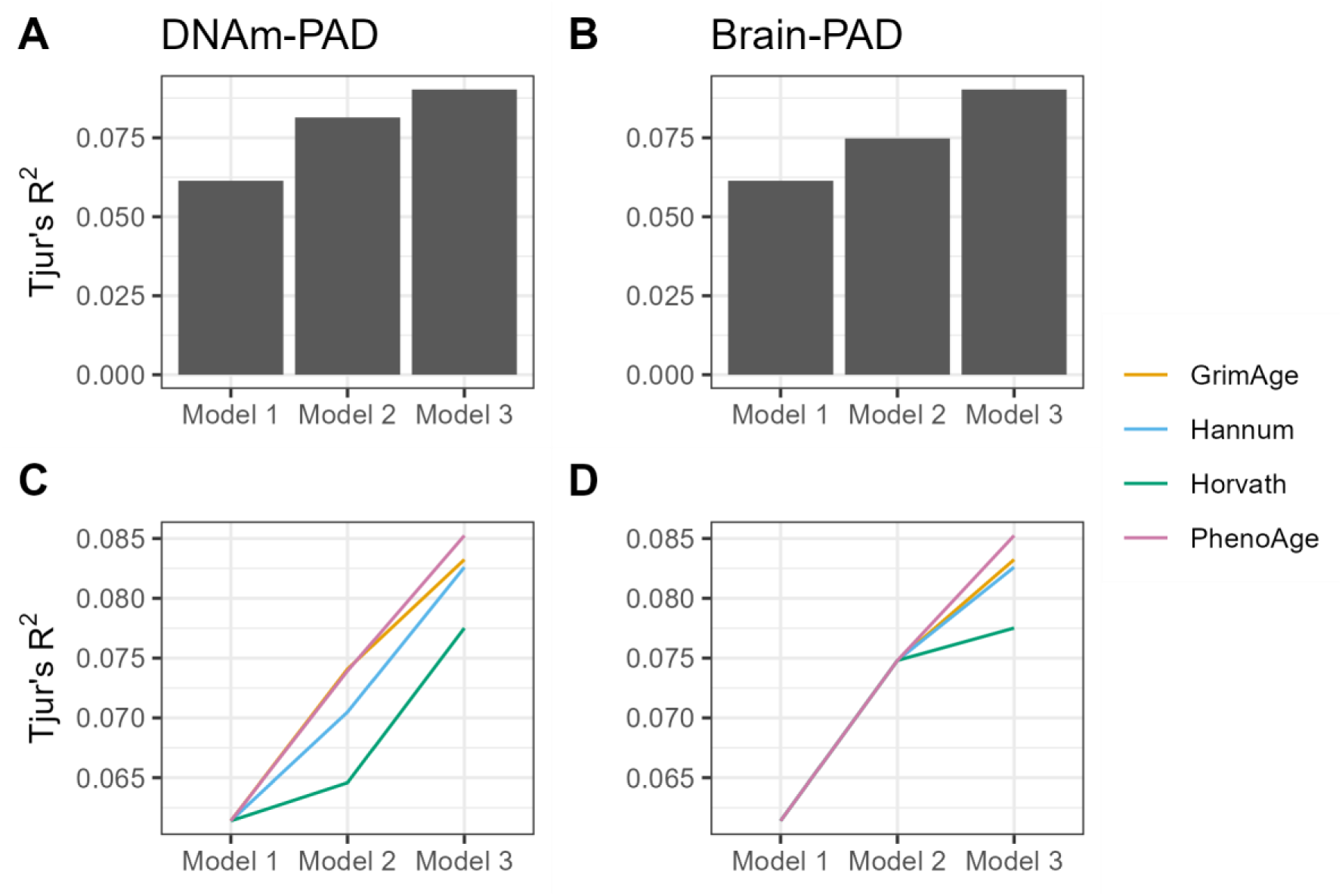
Comparisons of R^2^ across models of lifetime MDD in the Generation Scotland: Stratifying Resilience and Depression Longitudinally study. In panels A and B, all four DNAm-PAD measures are added simultaneously (Horvath, Hannum, PhenoAge and GrimAge); panels C and D present the increase in R^2^ for each DNAm-PAD measure individually when combined with Brain-PAD. Model 1 = Lifetime MDD predicted by age and sex only. Model 2 = Lifetime MDD predicted by age, sex and DNAm-PAD (Panel A) or Brain-PAD (Panel B). Model 3 = Lifetime MDD predicted by age, sex, DNAm-PAD and Brain-PAD.

### Inflammatory markers associated with MDD

Prior to testing mediation effects, we established a list of inflammatory markers associated with either lifetime MDD or current MDD. Regression models adjusting for age and sex showed associations between MDD phenotypes (particularly current MDD) and markers of peripheral inflammation, including white blood cells (WBC), neutrophils (NE), eosinophils (EO), monocytes (MO), C-reactive protein (CRP) and basophils (BA) (Table 2). A full table of all inflammatory markers and results are presented in Supplementary Table S2.

**Table 2:**
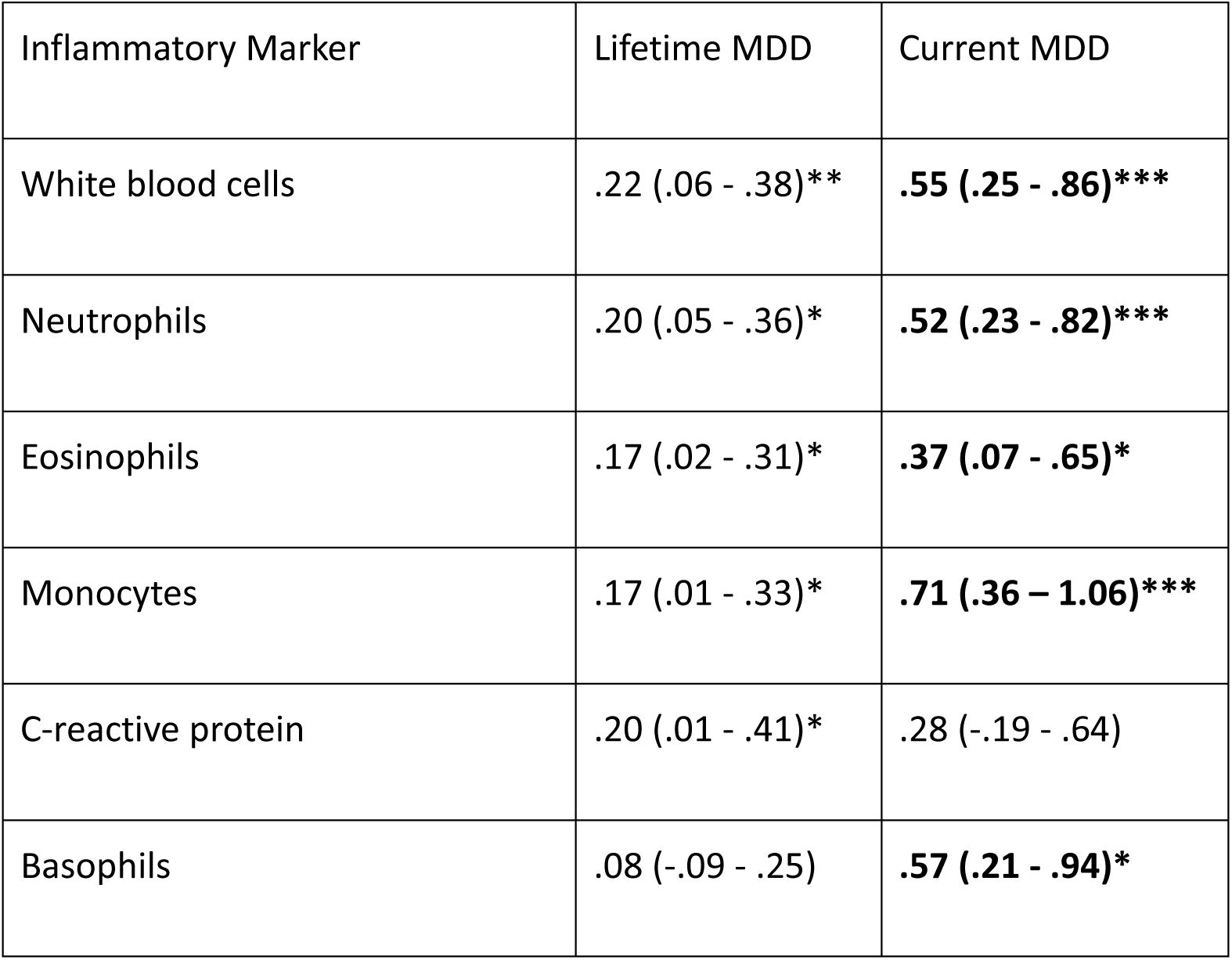
Inflammatory markers nominally associated with lifetime or current MDD in the Generation Scotland: Stratifying Resilience and Depression Longitudinally study. Log-transformed odds ratios (95% confidence intervals) are reported in the table. Significant associations surviving FDR correction for multiple testing are indicated in bold font. * = p<.05, ** = p<.01, *** = p<.001.

### Mediation effect of inflammatory markers on the association between BioAge-PAD and MDD

Mediation models for lifetime and current MDD were constructed using the six inflammatory markers identified above. While this was not the main focus of the current study, we also explored mediation effects for severe MDD and antidepressant use phenotypes for completeness. While no significant mediation effects of inflammatory markers were found for either the lifetime or current MDD phenotypes nor for severe MDD (Supplementary files S2-S4), there were significant mediation effects for the antidepressant use phenotype. WBC significantly mediated the associations between Brain-PAD and antidepressant use, and similarly for DNAm-PAD_GrimAge_. Additionally, both NE and MO significantly mediated the association between Brain-PAD and antidepressant use (see Supplementary Materials Table S3; Supplementary file S5).

## Discussion

This study explored (i) the relationship between markers of biological aging and MDD using different tissue types (DNAm and brain), (ii) their individual and combined associations with MDD, and (iii) potential mediation of the relationship between premature biological aging and MDD through inflammatory markers. These analyses were conducted in a relatively large population-based sample of unrelated individuals, with replication of brain age findings in an independent sample. We focused on two different but complementary types of biological aging: blood-based DNA methylation age and BrainAge. We also focused on two main MDD phenotypes, lifetime and current MDD. Full details of other MDD measures are described in the supplementary materials.

We found significant associations between lifetime MDD and DNAm-PAD_PhenoAge_, DNAm-PAD_GrimAge_ and Brain-PAD. The finding of increased brain aging was further validated in a large, independent sample from UK Biobank. This provides further demonstration of premature brain aging in MDD, across cohorts utilizing high-dimensional imaging data, adding to the literature on findings based on derive imaging phenotypes. Effect sizes of associations with MDD were comparable between Brain-PAD and DNAm-PADs (β_Brain-PAD_=.22; β_DNAm-PAD_ range=.20-.27) in GS:STRADL. We also report that a combination of blood- and brain-based measures of aging significantly improved classification of MDD cases and controls versus either measure in isolation in the context of MDD, consistent with similar studies on mortality(10), indicating separate and combined contributions of aging from DNAm-PAD and Brain-PAD measures. In particular, the combination of DNAm-PAD_PhenoAge_ and Brain-PAD explained the most variance associated with MDD. This study therefore provides evidence of premature biological aging in MDD, extending current literature through replication of increased brain aging in MDD and by bringing together different types of biological aging, demonstrating the importance of both peripheral and central biological aging processes in MDD.

Previous studies have typically investigated peripheral and neurobiological measures of biological aging separately in the context of MDD. Results for the second-generation DNAm clocks included in the present study – PhenoAge and GrimAge - have been largely mixed thus far. For example, some studies report positive associations between DNAmAge and MDD/depressive symptoms (19, 45–48) or no associations (49, 50). Additionally, these positive associations have been primarily reported for GrimAge(19, 45, 46). Only two previous studies have reported significant associations with PhenoAge in depression(51, 52). The first was a twin study which reported associations between PhenoAge and continuous depression scores (51). The second study examined relationships between childhood adversity and increased biological aging in individuals with depression(52). In terms of their derivation, GrimAge incorporates epigenetic markers of smoking, whereas PhenoAge incorporates phenotypic markers of aging including physiological status and morbidity profiles. Our results may therefore point to additional physiological aging processes in MDD that are captured by PhenoAge measures. Regarding Brain-Age, a recent meta-analysis reported a small increase in Brain-PAD in MDD (in the order of +0.90 (0.20) years), with some indication of variation based on whether Brain-PAD is calculated based on derived imaging metrics, as in ENIGMA mega-analysis studies, or whether it is derived based on the original high-dimensional imaging data, as in the current study(53). In sum, by leveraging concurrent neuroimaging and methylation data, our results point to premature biological aging across both the brain and periphery in MDD. We find separate and combined contributions of peripheral and central markers of aging, that are more predictive of MDD when combined than for either measure in isolation.

Finally, we also examined the contributions of inflammatory markers, first by testing associations with MDD phenotypes, then in mediation analyses between increased biological aging and MDD. We found the strongest relationship with inflammatory markers (from white blood cell counts) with current rather than lifetime MDD. This may suggest acute inflammatory processes are more strongly related to concurrent depressive symptom severity than for lifetime MDD as previously shown(27). White blood cells, which serve as a summary of multiple inflammatory markers, have been found previously to be associated with depressive symptoms, as supported by large-scale, cross-sectional (54) and longitudinal analyses (55). Contrary to our hypotheses, however, we did not find evidence for significant mediation of the association between increased biological aging and MDD through inflammatory markers. Before excluding the possibility of a role of inflammation in premature aging in MDD, future work could combine more stable and wider ranges biomarkers of inflammation, along with clinically ascertained samples, to robustly determine the importance of immune mediators of premature biological aging in MDD(26).

It is important to note as a limitation of the current study that the ethnic backgrounds of our samples were highly homogeneous (mainly Northern European), limiting the generalisability of our findings to other ethnic groups. We also note that the current study was cross-sectional and we did not examine directions of causality. This work could be expanded on using existing Mendelian Randomisation studies that indicate bidirectional relationships between aging and MDD(56, 57), to include different types of biological aging to gain a deeper understanding of its causal relationship with MDD.

In conclusion, our findings demonstrate that premature aging in MDD is observed across both brain and peripheral measures of biological aging, with evidence of shared and distinct contributions of each. While we did not find evidence of mediation of the relationship between premature aging and MDD through inflammatory markers, suggestions are made for future work to explore potential biological mechanisms in greater detail.

## Data Availability

All data produced in the present study are available upon reasonable request to the authors

## Author Contributions

EYX: Conceptualization, Methodology, Formal analysis, Investigation, Writing - Original Draft, Review and Editing, Visualization. CG: Writing - Original Draft, Review and Editing, DM: Methodology, Software, Writing - Review and Editing. LH: Writing - Review and Editing. KE: Data Curation, Writing - Review and Editing. RW: Data Curation, Writing - Review and Editing. DG: Data Curation, Writing - Review and Editing. DS: Data Curation, Writing - Review and Editing. GW: Data Curation, Writing - Review and Editing. AC: Data Curation, Writing - Review and Editing. SL: Supervision, Writing - Review and Editing. JC: Software, Writing - Review and Editing. AM: Supervision, Writing - Review and Editing. XS: Conceptualization, Methodology, Writing - Original Draft, Review and Editing, Supervision, Project administration. HW: Conceptualization, Methodology, Writing - Original Draft, Review and Editing, Supervision, Project administration.

## Abbreviations

BrainAge: biological age estimated from structural neuroimaging
Brain-PAD: predicted age difference between BrainAge and chronological age
BioAge: biological age
BioAge-PAD: predicted age difference between biological age and chronological age
DNAmAge: biological age estimated from DNA methylation, also known as epigenetic age
DNAm-PAD: predicted age difference between DNAmAge and chronological age
GS:STRADL: Generation Scotland: Stratifying Resilience and Depression
Longitudinally MDD: major depressive disorder
PAD: predicted age difference, i.e. difference between biological age and chronological age
UKB: UK Biobank

