## Supplementary Materials for "Epigenetic and Structural Brain Aging and their Associations with Major Depressive Disorder and Inflammatory Markers"

**DNAm age measures**

The "first-generation" DNAm clocks were trained on chronological age. Acceleration in DNAm Horvath and Hannum age – a DNAm age greater than chronological age – moderately predicted age-related morbidity (1-3). In contrast, "second-generation" clocks DNAmPhenoAge and DNAmGrimAge were developed using a two-stage approach (4, 5). These clocks were trained using a combination of clinical markers that associated with physical functioning and mortality (DNAmPhenoAge) and time-to-death (DNAmGrimAge). These included white blood cell count and C-reactive protein (for DNAmPhenoAge) and DNAm surrogates of leptin, adrenomedullin and smoking pack-years (for DNAmGrimAge), for example. In comparison to first-generation clocks, DNAmPhenoAge and DNAmGrimAge accelerations show stronger associations with mortality and time-to-death (4, 5). Details for each DNAm Age are listed below:

- *Horvath age:* An 'epigenetic clock' predicting age from DNA methylation at 353 CpG sites (2). During development, the model was trained on chronological age using methylation data from 8,000 samples taken from a range of tissues.
- *Hannum age:* An alternative epigenetic clock, also developed by training a model on chronological age and DNA methylation data. The training dataset for this model included more than 450,000 CpG sites, and 656 samples all taken from blood (3).
- *DNAmPhenoAge:* A more elaborate epigenetic clock trained not only on chronological age, but also on selected clinical factors associated with ageing-related mortality, such as cardiovascular disease, Alzheimer's disease, diabetes and chronic lower respiratory disease (4). This model predicts age from methylation at 513 CpG sites and was trained using the NHANES dataset.
- *DNAmGrimAge:* An epigenetic clock with a stronger focus on mortality and time-to-death rather than biological age itself. This was trained on DNA methylation estimates of seven plasma protein levels and DNAm-based estimates of smoking pack-years (5).

**Additional findings for severe MDD and antidepressant MDD case-control differences in BioAge-PAD**

**Severe MDD**

DNAm-PAD was significantly associated with severe MDD for $\text{DNAm-PAD}_{\text{PhenoAge}}$ ($\beta$ = .41; 95% CI: .05 - .77; p = .05) and $\text{DNAm-PAD}_{\text{GrimAge}}$ ($\beta$ = .52; 95% CI: .17 - .85; p < .01). Following FDR correction, $\text{DNAm-PAD}_{\text{GrimAge}}$ remained significantly associated with severe MDD. No significant associations were found for $\text{DNAm-PAD}_{\text{Horvath}}$, $\text{DNAm-PAD}_{\text{Hannum}}$ or Brain-PAD in both GS:STRADL and UKB.

**Antidepressant MDD**

Of the four DNAm-PAD measures, only DNAm-$PAD_{GrimAge}$ significantly associated with antidepressant use (β = .42; 95% CI: .12 - .72; p < .01). Brain-PAD also significantly associated with antidepressant use (β = .43, 95% CI: .18 - .68; p < .001). Consistent with the results from GS:STRADL, Brain-PAD in UKB was significantly associated with antidepressant use (β = .10; 95% CI: .02 - .18; p = .01). Both Brain-PAD and DNAm-$PAD_{GrimAge}$ associations survived FDR correction for multiple testing.

**Additive contributions of DNAm-PAD and Brain-PAD in severe MDD and antidepressant cases**

For severe MDD, the addition of DNAm-$PAD_{PhenoAge}$ improved model fit for Brain-PAD models ($\chi^2$ = 4.81; p = .03; ΔAUC = .04; $\Delta R^2$ = .01), as did DNAm-$PAD_{GrimAge}$ ($\chi^2$ = 8.16; p = .004; ΔAUC = .03; $\Delta R^2$ = .03).

For antidepressant use, the addition of Brain-PAD improved model fit for DNAm-$PAD_{Horvath}$ ($\chi^2$ = 4.00; p = .05; ΔAUC = .02; $\Delta R^2$ = .02) and DNAm-$PAD_{Hannum}$ ($\chi^2$ = 4.81; p = .03; ΔAUC = .04; $\Delta R^2$ = .01). Finally, the addition of DNAm-$PAD_{GrimAge}$ to the Brain-PAD model also improved model fit compared to Brain-PAD alone ($\chi^2$ = 5.58; p = .02; ΔAUC = .01; $\Delta R^2$ = .01).

**Figure S1: Scatter plots of chronological age plotted against predicted biological ages in GS:STRADL (A-E) and UK Biobank (F).**

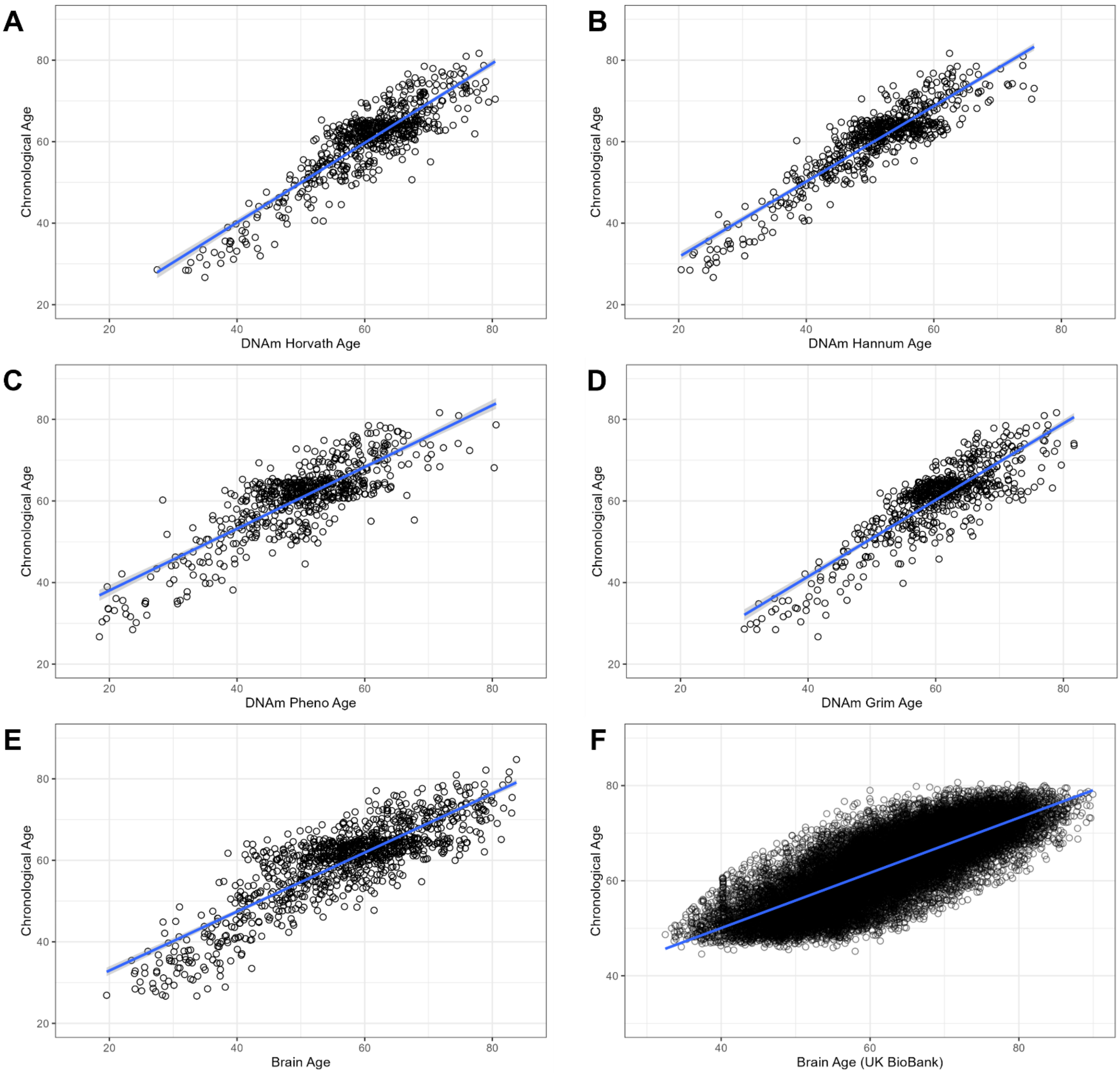


Figure 1: Scatter plots of chronological age plotted against predicted biological ages in GS:STRADL (A-E) and UK Biobank (F).

**Figure S2: GS:STRADL demographics by MDD phenotype**

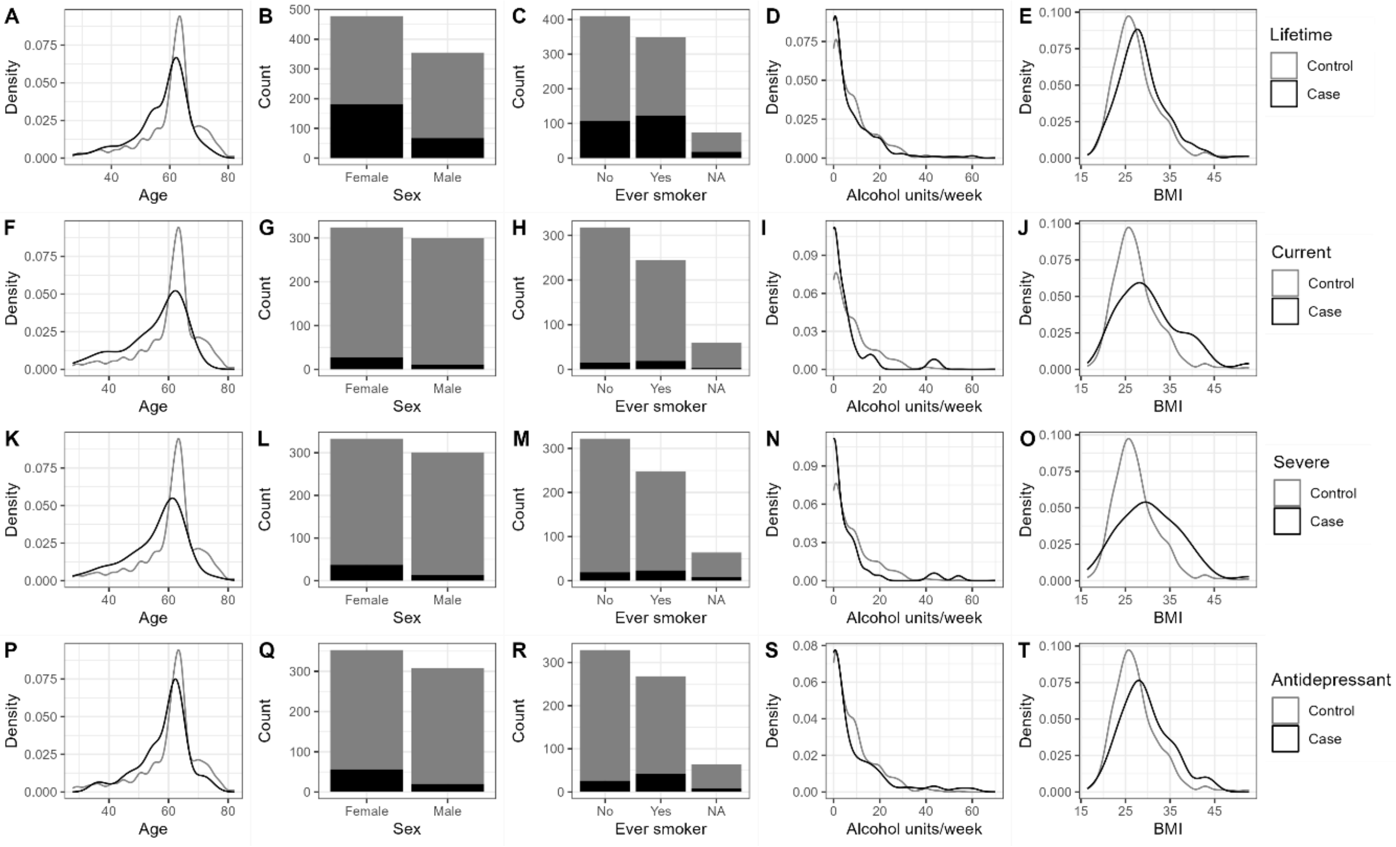


Figure S2: Demographic information for MDD phenotypes: lifetime (A-E), current (F-J), severe (K-O) and antidepressant use (P-T) in GS:STRADL. MDD cases are presented in black, MDD controls are presented in grey. BMI = body mass index.

**Figure S3: UK Biobank demographics by MDD phenotype**

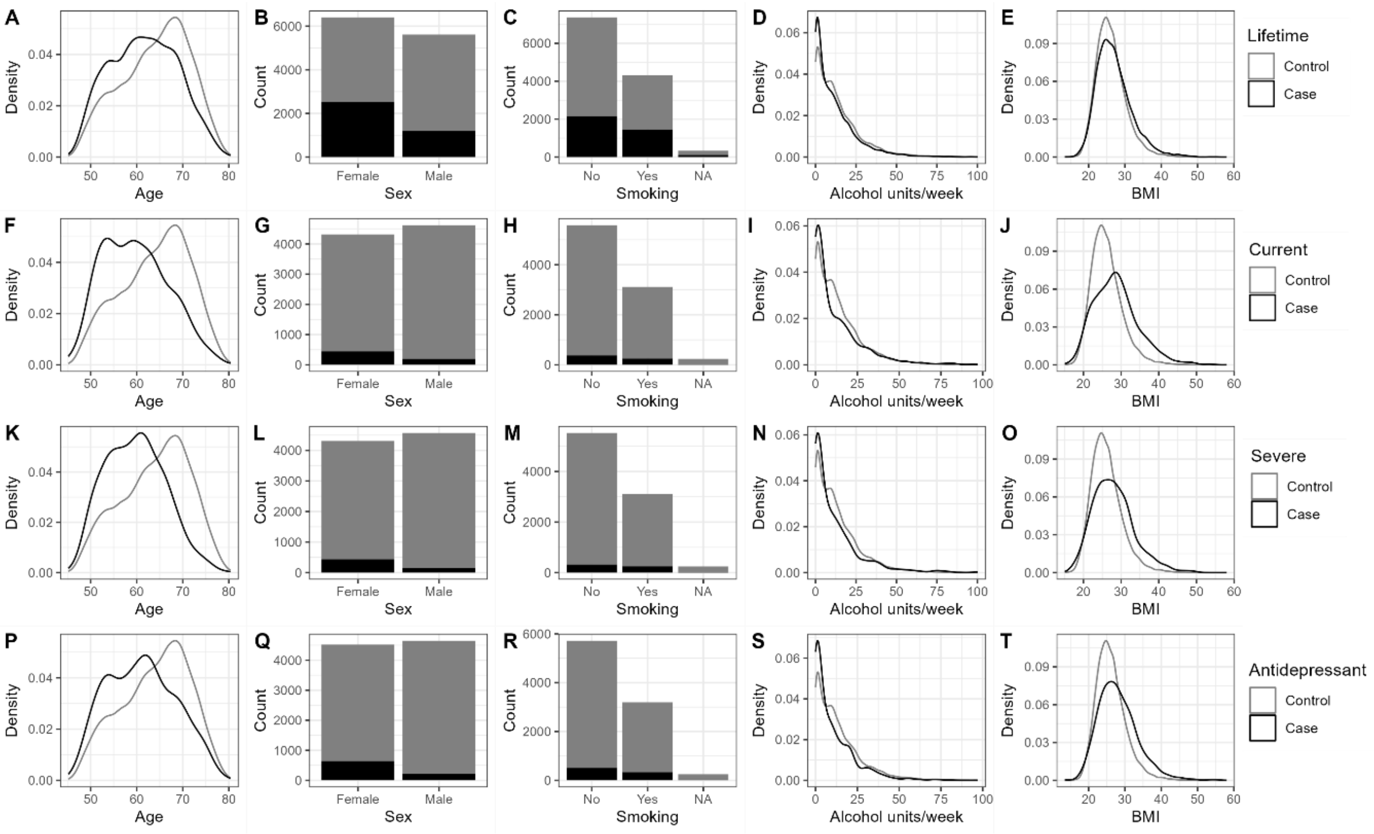


Figure S3: Demographic information for MDD phenotypes: lifetime (A-E), current (F-J), severe (K-O) and antidepressant use (P-T) in UK Biobank. MDD cases are presented in black, MDD controls are presented in grey. BMI = body mass index.

**Figure S4: Case-control differences in predicted biological age by MDD phenotype**

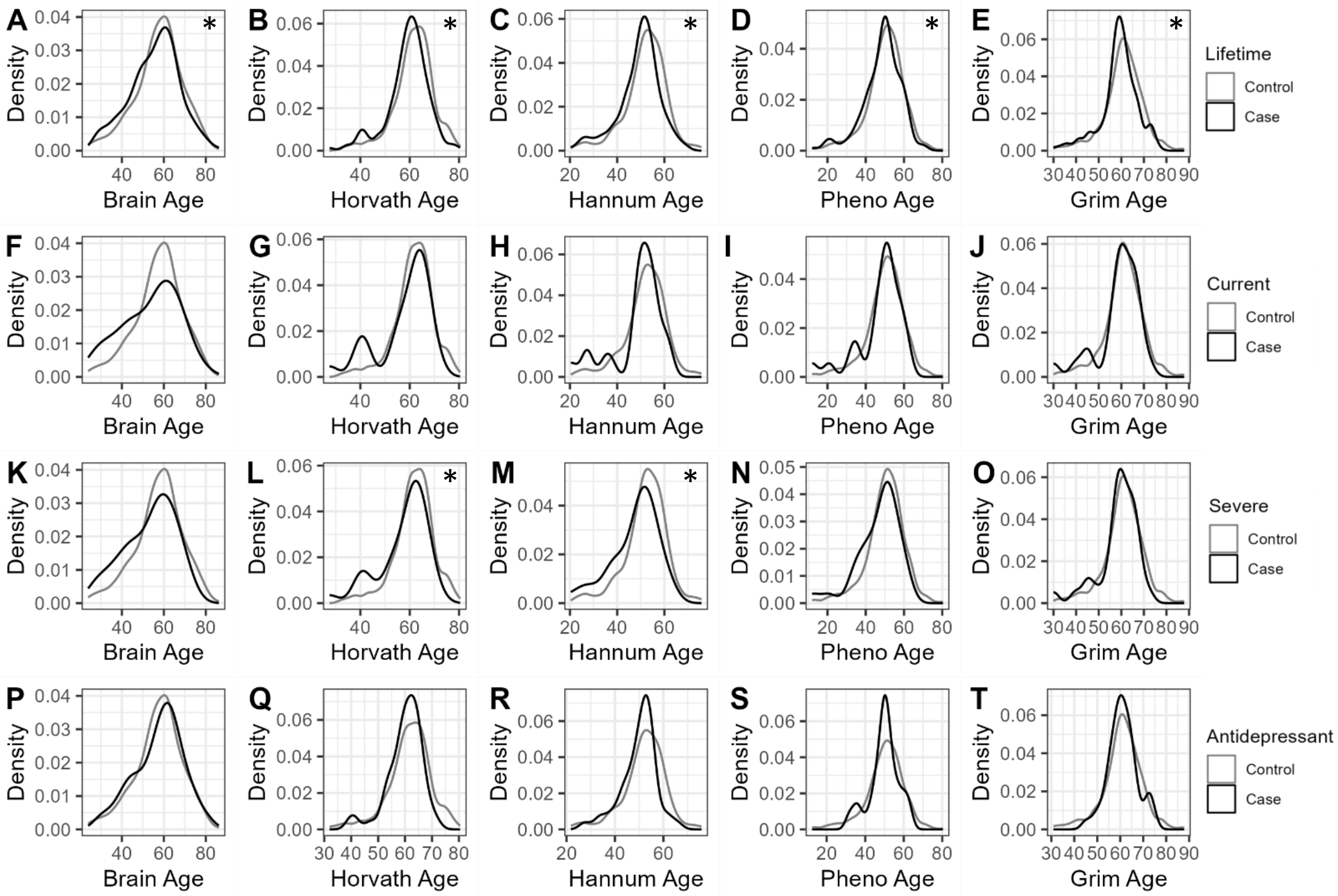


Figure S4: Distributions of predicted biological age for GS:STRADL cases (black) and controls (grey). *, ** and *** represent significant differences between mean case and control biological age at p<.05, p<.01 and p<.001, as determined by two-sample t-tests.

**Figure S5: Case-control differences in BioAge-PAD by MDD phenotype**

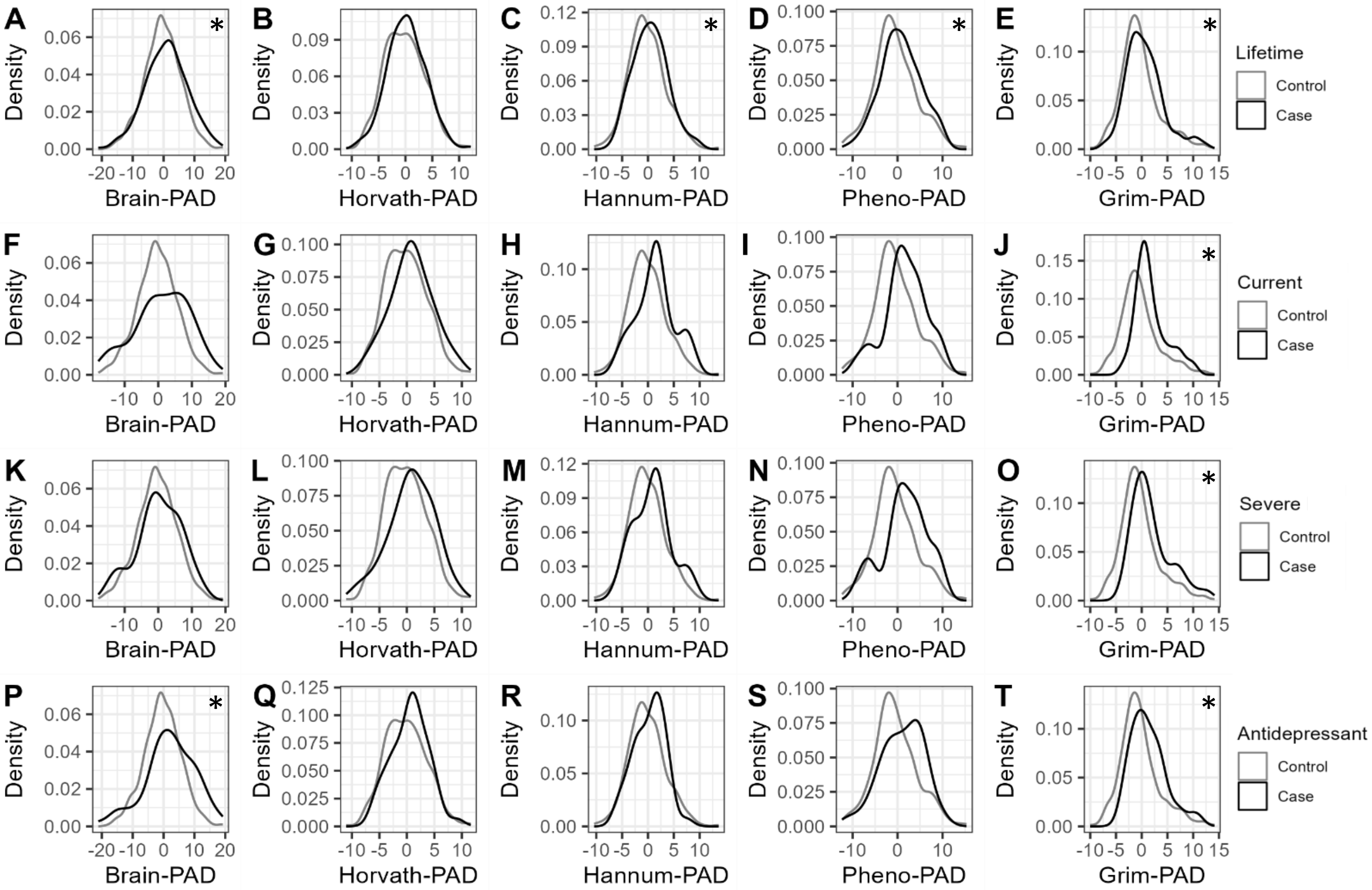


Figure S5: Distributions of biological age acceleration measures for GS:STRADL cases (black) and controls (grey). *, ** and *** represent significant differences between mean case and control biological age at p<.05, p<.01 and p<.001, as determined by two-sample t-tests. PAD = predicted age difference.

**Figure S6: Correlations between Bio-PAD measures in GS:STRADL**

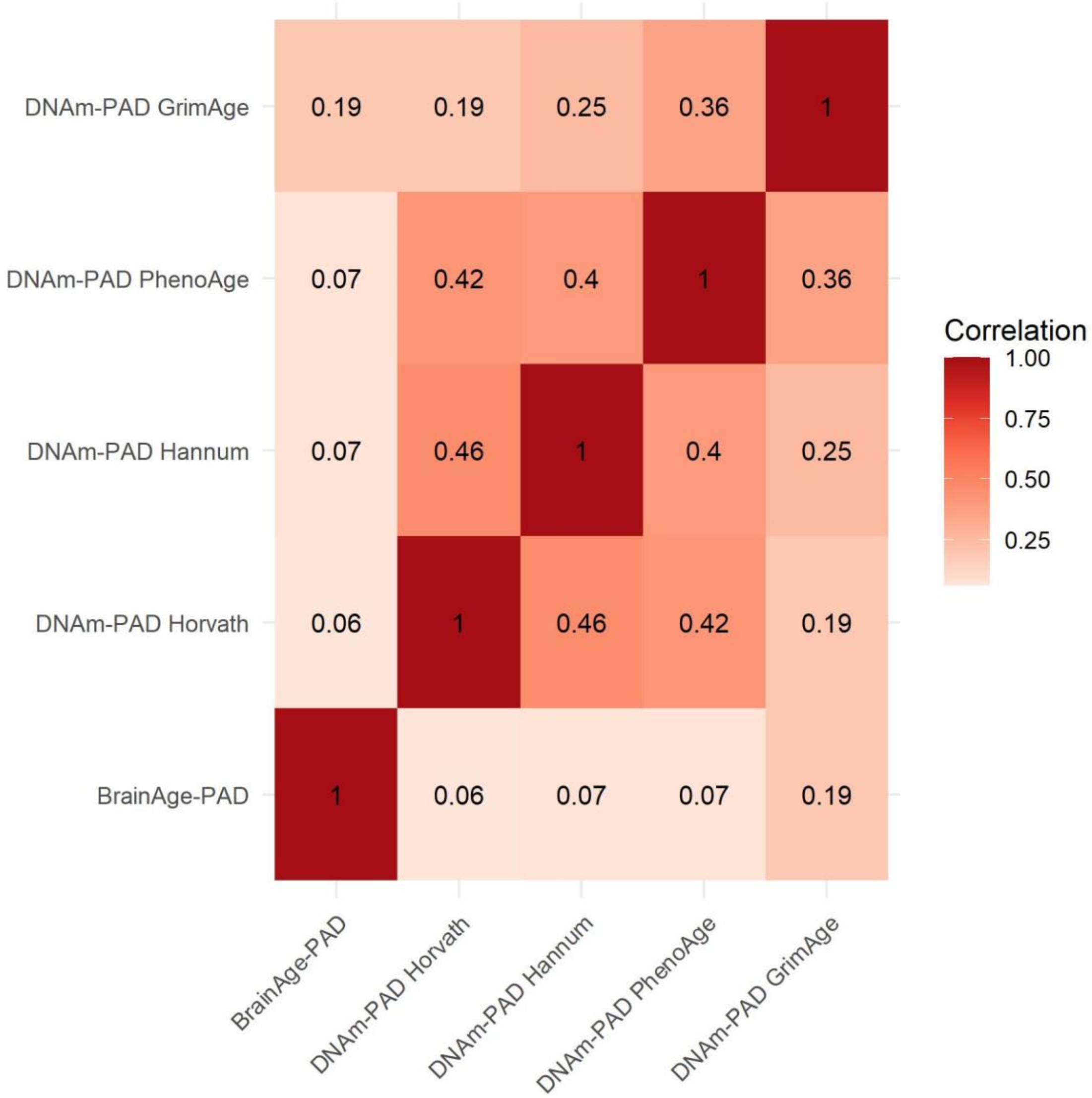


Figure S6: Heatmap of Pearson's correlations between BioAge-PAD measures in Generation Scotland: Stratifying Resilience and Depression Longitudinally. PAD = Predicted age difference.

**Table S1: MDD phenotypes within GS:STRADL and UK Biobank**

| | **GS:STRADL** | | **UK Biobank** | |
|---|---|---|---|---|
| | Lifetime MDD case (N = 248) | Control (N = 585) | Lifetime MDD case (N = 3717) | Control (N = 8301) |
| Current MDD [case % (N)] | 15.32% (38) | - | 17.24% (641) | - |
| Severe MDD [case % (N)] | 20.16% (50) | - | 15.58% (579) | - |
| Antidepressant use [case % (N)] | 30.65% (76) | - | 23.27% (865) | - |

Table S1: MDD phenotypes within Generation Scotland: Stratifying Resilience and Depression Longitudinally (GS:STRADL) and UK Biobank

**Table S2: Inflammatory markers associated with MDD phenotypes**

| Inflammatory Marker | Lifetime MDD | Current MDD | Severe MDD | Antidepressant use |
|---|---|---|---|---|
| White blood cell count (WBC) | .22 (.06 - .38)** | .55 (.25 - .86)*** | .50 (.23 - .78)*** | .54 (.30 - .80)*** |
| Neutrophil count (NE) | .20 (.05 - .36)* | .52 (.23 - .82)*** | .47 (.20 - .74)*** | .49 (.25 - .74)*** |
| Eosinophil count (EO) | .17 (.02 - .31)* | .37 (.07 - .65)* | .35 (.08 - .60)** | .18 (-.07 - .41) |
| Monocyte count (MO) | .17 (.01 - .33)* | .71 (.36 – 1.06)*** | .53 (.23 - .84)*** | .55 (.31 - .81)*** |
| C-reactive protein (CRP) | .20 (.01 - .41)* | .28 (-.19 - .64) | .41 (.11 - .70)** | .31 (.03 - .59)* |
| Basophil count (BA) | .08 (-.09 - .25) | .57 (.21 - .94)* | .60 (.29 - .92)*** | .46 (.20 - .72)*** |
| Haemoglobin (Hb) | .02 (-.15 - .20) | -.13 (-.38 - .21) | -.13 (-.37 - .16) | .06 (-.23 - .39) |
| Red blood cell count (RBC) | -.03 (-.20 - .14) | -.15 (-.41 - .17) | -.13 (-.38 - .16) | -.02 (-.29 - .29) |
| Haematocrit (HCT) | -.02 (NA - .12) | .004 (NA - .18) | -.01 (NA - .17) | -.02 (NA - .15) |
| Mean corpuscular volume (MCV) | .18 (-.003 - .39) | -.10 (-.28 - .17) | -.07 (-.25 - .18) | .43 (.06 - .81) * |
| Mean corpuscular haemoglobin (MCH) | .04 (-.10 - .21) | -.14 (-.34 - .10) | -.11 (-.31 - .11) | .18 (-.12 - .51) |
| Platelet count (PLT) | -.06 (-.23 - .10) | -.17 (-.57 - .21) | -.27 (-.62 - .07) | .18 (-.07 - .43) |
| Lymphocyte count (LY) | .13 (-.02 - .29) | .25 (-.08 - .58) | .27 (-.01 - .55) | .26 (.02 - .48) * |
| Large unstained cell count (LUC) | -.01 (-.19 - .14) | .02 (-.43 - .27) | .03 (-.33 - .26) | .09 (-.15 - .27) |
| Relative mean corpuscular | -.10 (-.26 - .04) | -.08 (-.27 - .28) | -.09 (-.27 - .21) | -.10 (-.27 - .12) |

| | | | | |
|---|---|---|---|---|
| haemoglobin count (MCHC_calc) | | | | |
| Neutrophil-lymphocyte ratio (NLR) | .06 (-.10 - .22) | .21 (-.16 - .52) | .21 (-.09 - .48) | .19 (-.06 - .41) |
| Lymphocyte-monocyte ratio (LMR) | -.03 (-.20 - .13) | -.39 (-.85 - .02) | -.16 (-.50 - .16) | -.27 (-.56 - .01) |
| Platelet-lymphocyte ratio (PLR) | -.16 (-.34 - .01) | -.45 (-.97 - .0004) | -.39 (-.80 - -.02) | -.13 (-.42 - .12) |
| Systemic inflammatory index (SII) | .03 (-.13 - .18) | .18 (-.16 - .46) | .15 (-.15 - .40) | .21 (-.01 - .42) |

Table S2: Standardised log-transformed odds ratios and 95% confidence intervals for each inflammatory blood marker.

*, ** and *** represent significant results at p<.05, p<.01 and p<.001, respectively.

**Table S3. Association between inflammatory markers and MDD phenotypes**

| Inflammatory Marker | Lifetime MDD | Current MDD | Severe MDD | Antidepressant use |
|---|---|---|---|---|
| WBC | .22 (.06 - .38)** | **.55 (.25 - .86)*** ** | **.50 (.23 - .78)*** ** | **.54 (.30 - .80)*** ** |
| NE | .20 (.05 - .36)* | **.52 (.23 - .82)*** ** | **.47 (.20 - .74)*** ** | **.49 (.25 - .74)*** ** |
| EO | .17 (.02 - .31)* | **.37 (.07 - .65)* ** | **.35 (.08 - .60)** ** | .18 (-.07 - .41) |
| MO | .17 (.01 - .33)* | **.71 (.36 – 1.06)*** ** | **.53 (.23 - .84)*** ** | **.55 (.31 - .81)*** ** |
| CRP | .20 (.01 - .41)* | .28 (-.19 - .64) | **.41 (.11 - .70)** ** | .31 (.03 - .59)* |

| | | | | |
|---|---|---|---|---|
| BA | .08 (-.09 - .25) | **.57 (.21 - .94)*** | **.60 (.29 - .92)***** | **.46 (.20 - .72)***** |

Table S3: Inflammatory markers associated with MDD phenotypes in GS:STRADL. Log-transformed odds ratios (95% confidence intervals) are reported in the table. WBC = white blood cells, NE = neutrophils, EO = eosinophils, MO = monocytes, CRP = C-reactive protein, BA = basophils.

Significant associations surviving FDR correction for multiple testing are indicated in bold font. * = p<.05, ** = p<.01, *** = p<.001.

**Table S4. Significant mediation effects of inflammatory markers for associations between BioAge-PAD and antidepressant use**

| BioAge-PAD | Mediator | Direct effect | Indirect effect | Total effect |
|---|---|---|---|---|
| Brain-PAD | WBC | .038 (.013 - .062)** | .006 (.001 - .011)* | .044 (.019 - .068)*** |
| DNAm-PAD GrimAge | WBC | .034 (.006 - .063)* | .007 (.001 - .013)* | .041 (.013 - .069)** |
| Brain-PAD | NE | .039 (.014 - .063)** | .005 (.0004 - .009)* | .044 (.019 - .068)*** |
| Brain-PAD | MO | .039 (.015 - .064)** | .004 (.0002 - .008)* | .044 (.019 - .068)*** |

Table S4: Direct effect (Bio-PAD -> MDD, controlling for inflammation), indirect effect (Bio-PAD -> inflammation -> MDD) and total effect (direct + indirect) for mediation models of bio-PAD, inflammation and antidepressant use. Log-transformed odds ratios (95% confidence intervals) are reported in the table. WBC = white blood cells, NE = neutrophils, EO = eosinophils, MO = monocytes, CRP = C-reactive protein, BA = basophils. * = unadjusted p<.05, ** = unadjusted p<.01, *** = unadjusted p<.001.
